# Multidimensional Social Vulnerability and Hepatic and Extrahepatic Outcomes in Adults With HIV/HBV Coinfection in the United States

**DOI:** 10.64898/2026.08.31.26361853

**Authors:** George A. Yendewa, Tayoot Chengsupanimit, Ali Dehghani Do, Ali Ahmed, Amir M. Mohareb, Chari Cohen, Michael L. Freeman, H. Nina Kim, Ighovwerha Ofotokun, Karine Dubé

**Author notes:** **Corresponding author:** George A. Yendewa, MD, MPH&TM, FACP, FIDSA Associate Professor of Medicine (Infectious Diseases) Case Reserve Western University School of Medicine Division of Infectious Diseases and HIV Medicine University Hospitals Cleveland Medical Center 11100 Euclid Ave, Cleveland, OH 44106, USA.

## Abstract

**Background:** HIV/HBV coinfection is associated with substantial liver-related morbidity and mortality, yet the impact of social vulnerability (SV) on clinical outcomes has not been systematically assessed. We evaluated associations of multidimensional SV with mortality, hepatic, virologic, and extrahepatic organ outcomes among adults with HIV/HBV.

**Methods:** We conducted a retrospective cohort study using TriNetX data from 110 U.S. healthcare organizations (2010-2026). We propensity score matched adults with HIV/HBV with and without documented SV 1:1 (2,024 per group). SV was defined using a four-domain framework encompassing material, healthcare access and engagement, interpersonal, and psychosocial vulnerability.

**Results:** Over 15,900 person-years, SV was associated with higher mortality (hazard ratio [HR], 2.06; 95% confidence interval [CI], 1.72-2.47), liver composite events (HR, 1.37; 95% CI, 1.07-1.76), hepatic decompensation (HR, 1.94; 95% CI, 1.39-2.70), hepatic failure (HR, 2.39; 95% CI, 1.53-3.73), HBV viremia (HR, 1.69; 95% CI, 1.32-2.16), and HIV viremia (HR, 2.05; 95% CI, 1.71-2.46). SV was also associated with major adverse cardiovascular events (HR, 1.47), chronic kidney disease (HR, 1.49), and diabetes (HR, 1.25). Multidomain SV generally showed stronger associations than single-domain SV for most hepatic and virologic outcomes, with HR ranges of 1.76-2.62 versus 1.35-1.76 for single-domain SV. Healthcare access and engagement vulnerability was most consistently associated with mortality and hepatic outcomes.

**Conclusions:** SV was associated with mortality, hepatic disease, impaired HIV/HBV control, extrahepatic organ morbidity, and acute care utilization in adults with HIV/HBV. SV assessment may improve risk stratification and identify actionable intervention targets during HIV/HBV care.

**Graphical abstract:** 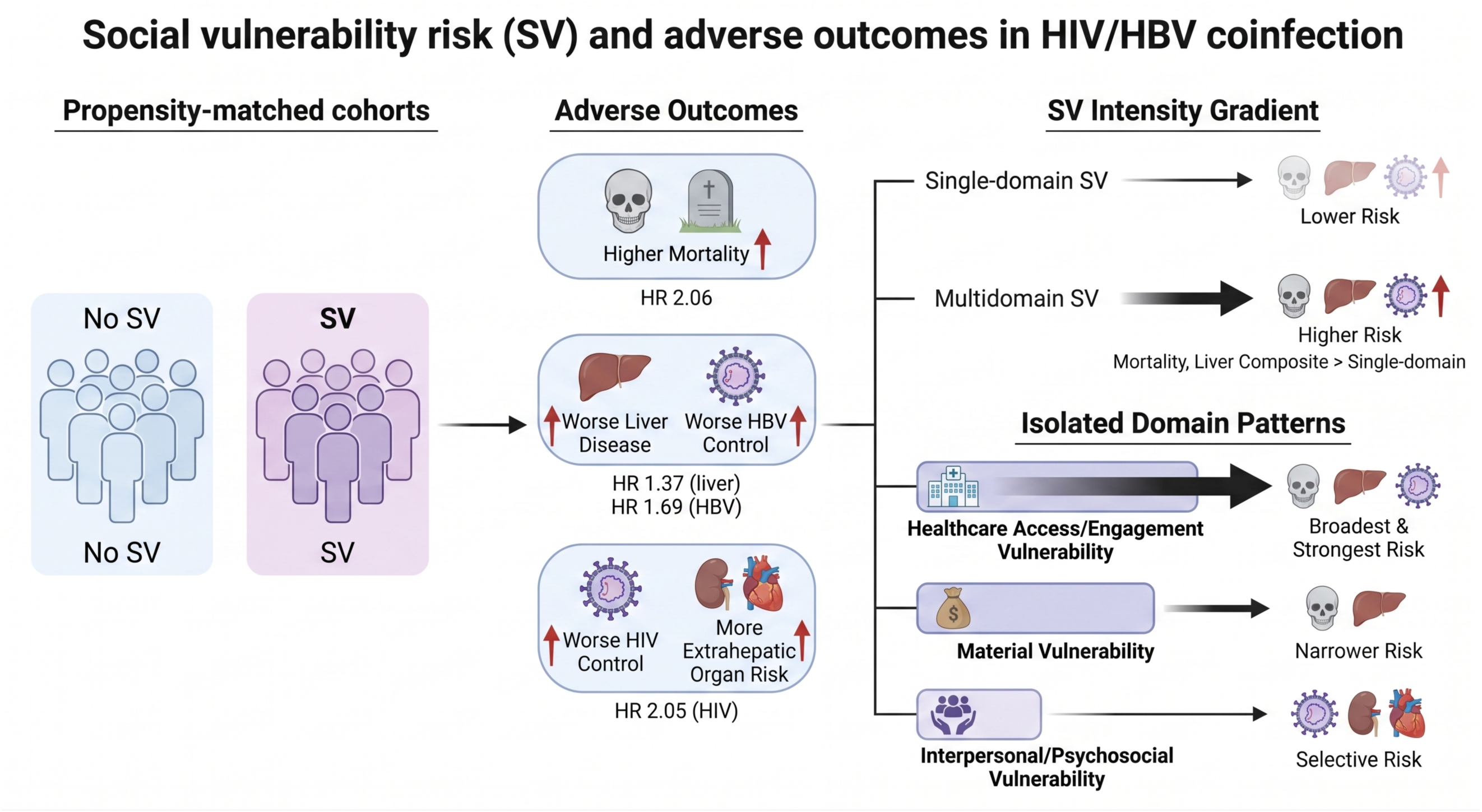

## INTRODUCTION

Human immunodeficiency virus (HIV) and hepatitis B virus (HBV) coinfection remains a clinically consequential dual viral condition [1,2]. Although HBV-active antiretroviral therapy (ART) has improved survival and reduced liver-related morbidity, people with HIV/HBV remain at elevated risk for persistent HBV replication, progressive liver disease, hepatocellular carcinoma (HCC), impaired immune recovery, and death [1–6]. HIV/HBV is also increasingly recognized as a multisystem condition in which hepatic risk may coexist with cardiovascular disease, chronic kidney disease (CKD), and metabolic disease [7–12]. Understanding the social and structural factors that shape this clinical phenotype is essential for improving long-term outcomes.

Social determinants of health (SDoH) are recognized drivers of HIV outcomes. Among people with HIV, material hardship, incarceration, healthcare barriers, and interpersonal violence are associated with delayed diagnosis, poor retention in care, lower ART adherence, viral nonsuppression, and mortality [13–16]. These dimensions of social vulnerability (SV) may be especially relevant to HIV/HBV coinfection, where successful long-term outcomes depend on uninterrupted HBV-active ART, sustained HIV and HBV viral control, regular liver disease monitoring, and HCC surveillance [1,17,21,22]. Food insecurity and interpersonal violence have been linked to immune activation, liver fibrosis, HIV nonsuppression, and adverse clinical outcomes [18–20], while insurance gaps, missed visits, and fragmented healthcare access may disrupt viral suppression and monitoring continuity [17,21,22].

Yet, SV is often modeled as a static background condition or examined as isolated social risk indicators. This approach may miss the multidimensional and dynamic ways in which social adversity shapes disease trajectories, especially in syndemic conditions such as HIV/HBV coinfection [23–29]. In prior work, we proposed a dynamic SV framework for HIV/HBV coinfection, conceptualizing SV as a time-varying and potentially actionable risk phenotype across four clinically recognized domains: material vulnerability, healthcare access and engagement, interpersonal adversity, and psychosocial vulnerability [26]. This framework allows SV to be evaluated by presence, intensity, and domain involvement.

In this study, we applied this multidimensional framework to a large U.S. matched cohort of adults with HIV/HBV coinfection. We examined associations between documented SV and mortality, hepatic outcomes, HIV and HBV disease control, and extrahepatic organ outcomes. We also evaluated whether clinical risk varied by SV intensity and isolated SV domain involvement, and whether associations persisted after accounting for baseline healthcare utilization.

## METHODS

### Data Source

We conducted a retrospective cohort study using the TriNetX Research Network, a federated, deidentified electronic health record platform containing structured patient-level data from participating healthcare organizations, including demographics, diagnoses, procedures, medications, laboratory values, vital status, and healthcare utilization. We analyzed data from 110 healthcare organizations in the United States. TriNetX has obtained institutional review board exemption from the Western Institutional Review Board; because we used only deidentified data, additional institutional review board approval was not required for this study.

### Cohort Definition, Index Date, and Follow-up

We included adults aged ≥18 years with qualifying events on or after January 1, 2010, and ≥6 months of electronic health record activity before index. We defined HIV using relevant diagnosis codes. We defined chronic HBV infection using chronic hepatitis B diagnosis codes, positive hepatitis B surface antigen, or detectable HBV DNA documented ≥6 months apart. The analytic population included adults with evidence of both HIV and chronic HBV before cohort entry. We excluded individuals with hepatitis C virus infection or prior liver transplantation to reduce outcome misclassification. The index date was the date each individual first met cohort criteria, defined separately for the SV and no-SV cohorts. Outcome follow-up began 1 day after index and continued until outcome occurrence, censoring, or the end of available data. Complete code lists are provided in the Supplementary File.

### Social Vulnerability Exposure

We defined the primary exposure as documented SV, identified by ≥1 qualifying social risk, social circumstance, or maltreatment-related code at or before cohort entry. The comparator group included adults with HIV/HBV coinfection and no documented SV codes from the prespecified code set. We operationalized SV using our four-domain framework: material vulnerability, healthcare access and engagement vulnerability, interpersonal adversity, and psychosocial vulnerability (**Figure 1**).

**Figure 1.**
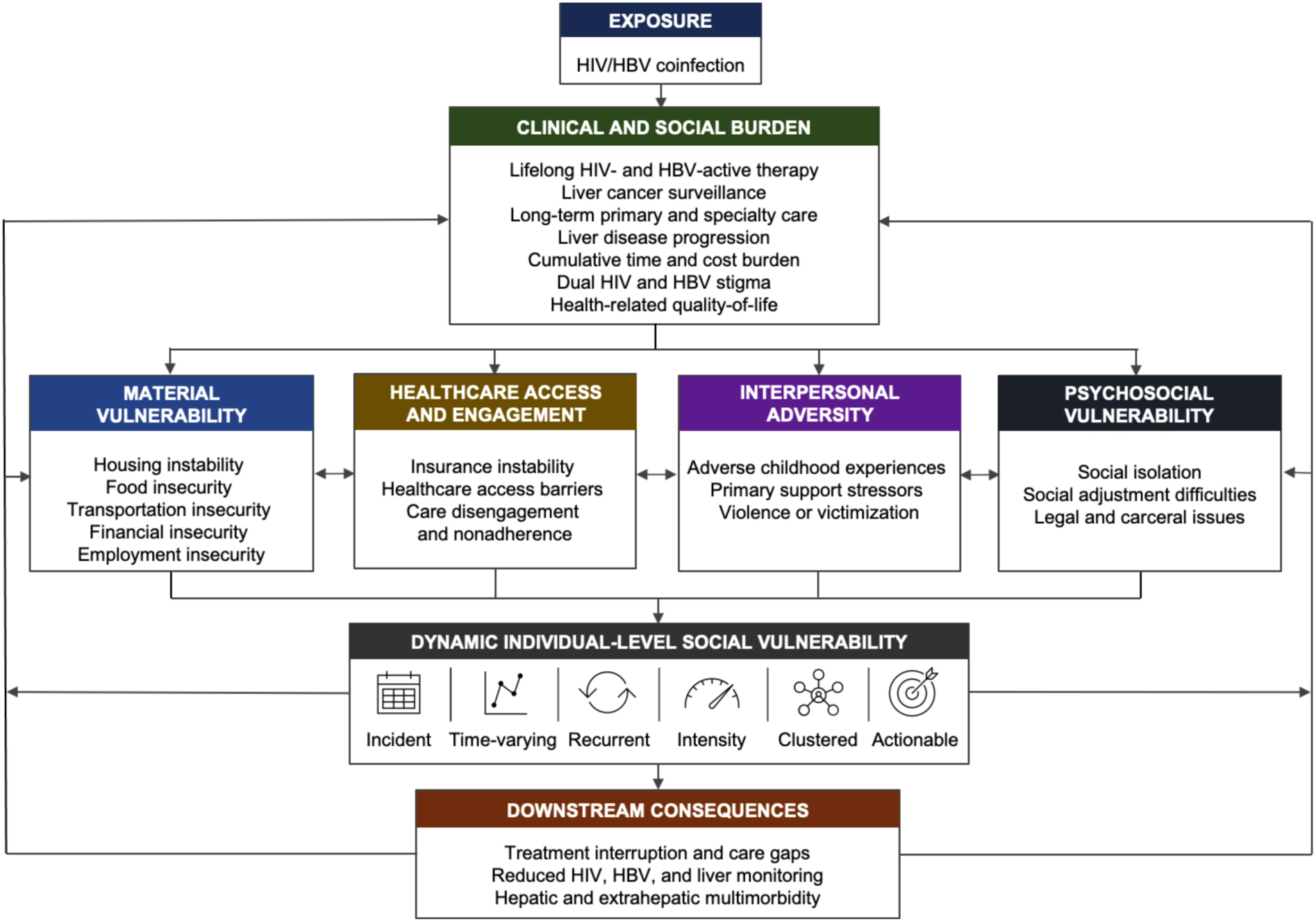
Conceptual Framework for Dynamic Individual-Level Social Vulnerability in HIV/HBV Coinfection HIV/HBV coinfection is conceptualized as an exposure that may generate clinical and social burden through lifelong HIV- and HBV-active therapy, HCC surveillance, long-term primary and specialty care, dual HIV and HBV stigma, disease progression, and health-related quality-of-life burden. These burdens may contribute to four interrelated domains of social vulnerability: material vulnerability, healthcare access and engagement, interpersonal adversity, and social and environmental vulnerability. Collectively, these domains define a dynamic individual-level social vulnerability phenotype characterized by incident onset, time variation, recurrence, intensity, clustering, and clinical actionability. Potential downstream consequences include treatment interruption and care disengagement, reduced viral and liver disease monitoring continuity, and hepatic and extrahepatic multimorbidity.

Material vulnerability included housing instability, food insecurity, economic or employment insecurity, transportation insecurity, and related material hardship. Healthcare access and engagement vulnerability included barriers to health system access, loss of insurance, treatment nonadherence, and care disengagement. Interpersonal adversity included physical or sexual abuse, neglect, maltreatment, violence, and other interpersonal safety threats. Psychosocial vulnerability captured broader problems related to primary support, social environment, and related social-contextual stressors.

In the primary analyses, we compared individuals with any documented SV with those with no documented SV. In secondary analyses, we evaluated two prespecified dimensions of the framework: SV intensity and isolated SV domain involvement. For intensity analyses, we defined single-domain vulnerability as exactly 1 affected domain and multidomain vulnerability as ≥2 affected domains. For isolated-domain analyses, we compared individuals with exactly 1 affected domain and no other documented domains with individuals with no documented SV. Because of limited sample size, we combined interpersonal and psychosocial vulnerability domains in isolated-domain analyses.

### Outcomes

We selected outcomes based on the clinical consequences of HIV/HBV coinfection and the hypothesized pathways linking SV to liver disease, viral control, and extrahepatic organ morbidity [1,17,29]. The primary outcomes were all-cause mortality and newly documented hepatic and HBV virologic outcomes [30,31]. These included a liver composite and its individual components: cirrhosis, hepatic decompensation, hepatic failure, and HCC [30,31]. Hepatic decompensation was defined as ascites, esophageal varices, spontaneous bacterial peritonitis, hepatic encephalopathy, or hepatorenal syndrome. HBV viremia was defined as HBV DNA ≥2,000 IU/mL. Liver inflammation outcomes included significant alanine aminotransferase (ALT) elevation and significant aspartate aminotransferase (AST) elevation, each defined as ≥3 times the upper limit of normal or ≥100 U/L. Liver synthetic function outcomes included thrombocytopenia, hypoalbuminemia, and coagulopathy.

Secondary clinical outcomes included HIV disease control and extrahepatic organ outcomes. HIV disease control outcomes included HIV RNA >50 copies/mL and CD4 count <200 cells/µL. Extrahepatic organ outcomes included cardiovascular disease, CKD, and diabetes. Cardiovascular outcomes included major adverse cardiovascular events (MACE), ischemic heart disease, heart failure, and cerebrovascular disease. MACE was defined as a composite of cardiovascular or undetermined-cause death, cardiac arrest, myocardial infarction or ischemic heart disease, heart failure, cerebrovascular or transient ischemic events, peripheral vascular disease, and coronary or peripheral revascularization [32]. CKD was defined using diagnosis codes, dialysis-related procedure codes, or estimated glomerular filtration rate-based laboratory criteria. Diabetes was defined using diagnosis codes, hemoglobin A1c ≥6.5%, or receipt of antidiabetic medication. Exploratory laboratory outcomes, including C-reactive protein (CRP) and erythrocyte sedimentation rate (ESR), are reported in the **Supplementary File**.

### Covariates and Propensity Score Matching

We performed 1:1 propensity score matching without replacement to reduce baseline differences between adults with HIV/HBV coinfection with and without documented SV. We selected covariates based on their potential association with SV documentation and clinical outcomes. The primary matching model included demographics, comorbidities, and substance-related risk factors. We used baseline covariates that overlapped with outcome domains to balance pre-index risk; corresponding outcome analyses excluded individuals with pre-window documentation of that outcome.

### Sensitivity Analyses

We conducted three prespecified sensitivity analyses. Because greater healthcare contact may increase opportunities for both SV documentation and outcome ascertainment, we designated healthcare utilization adjustment as the key sensitivity analysis. This analysis accounted for ambulatory visits, emergency department visits, and hospitalizations before index to evaluate whether associations persisted after accounting for differential baseline healthcare contact. We then conducted two additional sensitivity analyses. First, we applied a 6-month lag, starting follow-up 6 months after index, to reduce misclassification from conditions present before cohort entry but first documented during early evaluation. Second, we separately applied propensity score matching that included CD4 count, HIV RNA, HBV DNA, and ART to account for baseline disease severity, viral activity, and treatment exposure.

### Statistical Analysis

We used the TriNetX Advanced Analytics platform to perform all analyses. We summarized continuous variables as means with standard deviations or medians with interquartile ranges and categorical variables as frequencies and percentages. We calculated incidence rates per 100 person-years with 95% confidence intervals (CIs). We estimated hazard ratios (HRs) with 95% CIs using Cox proportional hazards models. We censored individuals after outcome occurrence, end of electronic health record activity, or end of available data. We considered P values <0.05 statistically significant.

## RESULTS

### Baseline Characteristics

Before matching, 2,188 adults with HIV/HBV coinfection had ≥1 documented SV indicator and 7,769 had none (**Table 1**). Age was similar between groups, but the SV group had higher proportions of women (24.5% vs. 19.7%) and Black or African American individuals (55.4% vs. 39.1%), and fewer Asian individuals (1.6% vs. 5.0%). Comorbidities were more prevalent in the SV group, including cardiovascular diseases, diabetes, chronic lower respiratory diseases, CKD, liver disease including cirrhosis, and neoplasms. Mental health diagnoses and substance use disorders were also more common in the SV group before matching. After 1:1 propensity score matching, 2,025 adults with SV were matched to 2,025 without SV. The matched cohorts were well balanced across measured baseline characteristics. Additional baseline HIV and HBV indices, ART exposure, laboratory availability, and healthcare utilization characteristics are shown in **Supplementary Table S1**.

**Table 1.**
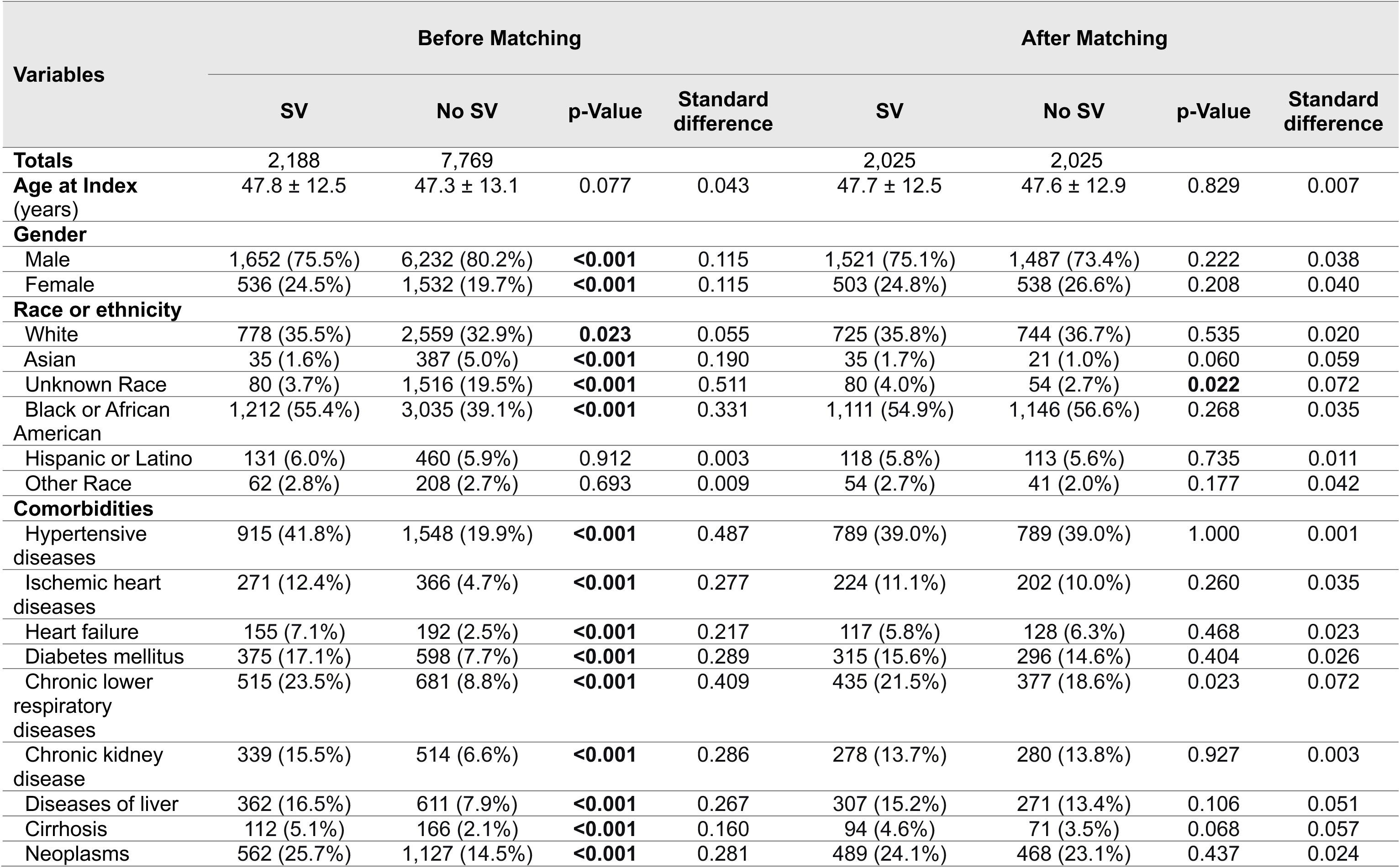
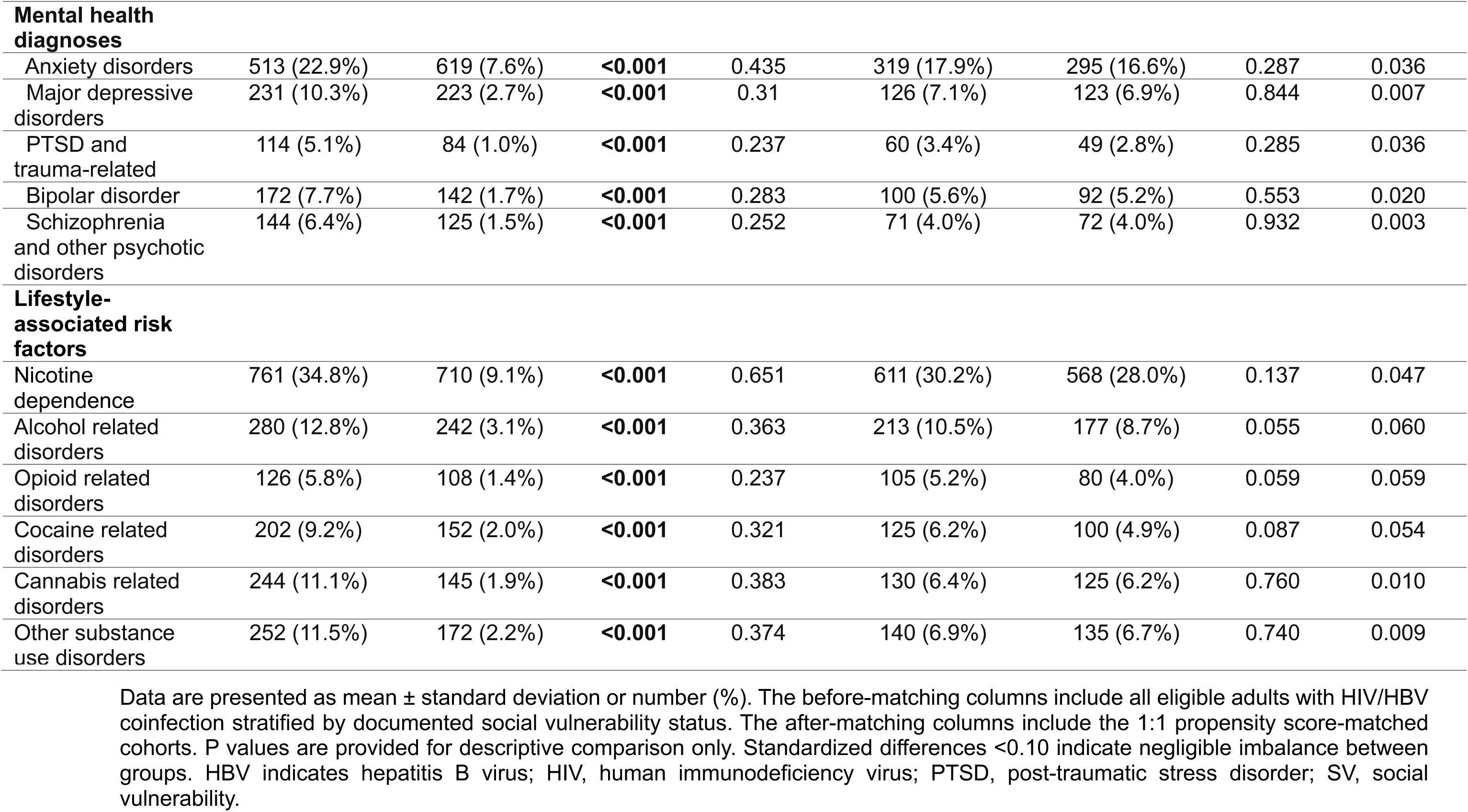
Baseline Characteristics of Adults With HIV/HBV Coinfection With and Without Documented Social Vulnerability Before and After Propensity Score Matching.

### Matched Analysis of Mortality, Hepatic, Virologic, and Extrahepatic Organ Outcomes

Over 15,900 person-years of observation, SV was associated with higher all-cause mortality (HR, 2.06; 95% CI, 1.72-2.47), with incidence rates of 4.7 and 2.1 per 100 person-years in the SV and no-SV groups, respectively (**Table 2**). SV was associated with a higher risk of the liver composite (HR, 1.37; 95% CI, 1.07-1.76), hepatic decompensation (HR, 1.94; 95% CI, 1.39-2.70), and hepatic failure (HR, 2.39; 95% CI, 1.53-3.73), while cirrhosis and HCC were not significantly different. SV was also associated with HBV viremia (HR, 1.69; 95% CI, 1.32-2.16), significant ALT elevation (HR, 1.32; 95% CI, 1.08-1.62), significant AST elevation (HR, 1.50; 95% CI, 1.26-1.79), and markers of impaired liver synthetic function, including thrombocytopenia, hypoalbuminemia, and coagulopathy (HR range, 1.29-1.48). Complete incidence rates and hazard ratios, including exploratory inflammatory markers, are shown in **Supplementary Table S2**.

**Table 2.** Incidence Rates and Hazard Ratios for Mortality, Hepatic, Virologic, and Multisystem Outcomes by Social Vulnerability Status in Propensity Score-Matched Adults With HIV/HBV Coinfection.

| Outcomes | Cohorts |  |  | IR (95% CI)<br>(per 100 person-years) |  | HR (95% CI) | p-Value |
| --- | --- | --- | --- | --- | --- | --- | --- |
|  | At risk | SV | No SV | SV | No SV |  |  |
| **Follow up, Mean ± SD (years)** | 3.95 ± 3.72 | 3.34 ± 3.37 | 4.57 ± 4.06 | | | | |
| **Follow up, Median (IQR) (years)** | 2.86 (5.25) | 2.18 (4.32) | 3.54 (6.17) |  |  |  |  |
| **Survival** |  |  |  |  |  |  |  |
| All-cause mortality | 4,031 | 316 (15.7%) | 195 (9.7%) | 4.7 (4.2-5.2) | 2.1 (1.8-2.4) | 2.06 (1.72-2.47) | **<0.001** |
| **Liver disease** |  |  |  |  |  |  |  |
| Liver composite | 3,593 | 131 (7.4%) | 126 (6.9%) | 2.2 (1.9-2.6) | 1.5 (1.3-1.8) | 1.37 (1.07-1.76) | **0.011** |
| Cirrhosis | 3,765 | 75 (4.0%) | 84 (4.4%) | 1.2 (1.0-1.5) | 1.0 (0.8-1.2) | 1.21 (0.88-1.65) | 0.244 |
| Hepatic decompensation | 3,828 | 89 (4.7%) | 59 (3.0%) | 1.4 (1.1-1.7) | 0.7 (0.5-0.9) | 1.94 (1.39-2.70) | **<0.001** |
| Hepatic failure | 3,910 | 56 (2.9%) | 30 (1.5%) | 0.9 (0.7-1.1) | 0.3 (0.2-0.5) | 2.39 (1.53-3.73) | **<0.001** |
| HCC | 3,992 | 16 (0.8%) | 19 (1.0%) | 0.2 (0.1-0.4) | 0.2 (0.1-0.3) | 1.04 (0.53-2.03) | 0.905 |
| **HBV viremia and liver inflammation** |  |  |  |  |  |  |  |
| HBV DNA ≥2,000 IU/mL | 3,782 | 151 (8.2%) | 114 (5.9%) | 2.5 (2.1-2.9) | 1.3 (1.1-1.6) | 1.69 (1.32-2.16) | **<0.001** |
| Significant ALT elevation (≥100 IU/L) | 2,898 | 186 (13.5%) | 192 (12.7%) | 4.0 (3.5-4.7) | 2.8 (2.4-3.2) | 1.32 (1.08-1.62) | **0.007** |
| Significant AST elevation (≥100 IU/L) | 2,407 | 239 (22.9%) | 263 (19.3%) | 6.9 (6.0-7.8) | 4.2 (3.7-4.8) | 1.50 (1.26-1.79) | **<0.001** |
| **Liver synthetic function** |  |  |  |  |  |  |  |
| Thrombocytopenia (platelet count $<150 \times 10^9$/L) | 2,397 | 204 (18.9%) | 233 (17.7%) | 5.7 (4.9-6.5) | 3.9 (3.4-4.4) | 1.29 (1.07-1.56) | **0.008** |
| Hypoalbuminemia (albumin $<3.5$ g/dL) | 2,236 | 234 (24.9%) | 273 (21.1%) | 7.5 (6.6-8.5) | 4.6 (4.1-5.2) | 1.48 (1.24-1.77) | **<0.001** |
| Coagulopathy (INR $>1.5$) | 3,308 | 111 (6.9%) | 104 (6.2%) | 2.0 (1.7-2.4) | 1.4 (1.2-1.7) | 1.35 (1.03-1.76) | **0.024** |
| **HIV disease control** |  |  |  |  |  |  |  |
| HIV RNA $>50$ copies/mL | 2,370 | 243 (25.0%) | 234 (16.7%) | 7.5 (6.6-8.5) | 3.7 (3.2-4.2) | 2.05 (1.71-2.46) | **<0.001** |
| CD4 count <200 cells/μL | 2,647 | 172 (15.3%) | 177 (11.6%) | 4.6 (3.9-5.3) | 2.5 (2.2-2.9) | 1.58 (1.28-1.95) | **<0.001** |
| **Extrahepatic outcomes** |  |  |  |  |  |  |  |
| MACE | 3,092 | 224 (15.3%) | 222 (13.7%) | 4.6 (4.0-5.2) | 3.0 (2.6-3.4) | 1.47 (1.22-1.77) | **<0.001** |
| Ischemic heart disease | 3,238 | 166 (10.7%) | 146 (8.6%) | 3.2 (2.7-3.7) | 1.9 (1.6-2.2) | 1.61 (1.29-2.02) | **<0.001** |
| Heart failure | 3,435 | 126 (7.6%) | 86 (4.9%) | 2.3 (1.9-2.7) | 1.1 (0.9-1.4) | 1.99 (1.51-2.62) | **<0.001** |
| Cerebrovascular disease | 3,443 | 139 (8.3%) | 112 (6.4%) | 2.5 (2.1-2.9) | 1.4 (1.2-1.7) | 1.67 (1.30-2.15) | **<0.001** |
| CKD | 2,885 | 164 (12.2%) | 158 (10.2%) | 3.7 (3.1-4.3) | 2.2 (1.9-2.6) | 1.49 (1.20-1.86) | **<0.001** |
| Diabetes | 2,816 | 234 (17.2%) | 258 (17.7%) | 5.2 (4.6-5.9) | 3.9 (3.4-4.4) | 1.25 (1.04-1.49) | **0.015** |

Hazard ratios compare adults with HIV/HBV coinfection with versus without documented social vulnerability after propensity score matching. Outcome-specific analytic cohort, n, indicates the number at risk for each outcome after exclusion of individuals with pre-follow-up evidence of that outcome; therefore, denominators vary across rows. Incidence rates are per 100 person-years. Significant ALT and AST elevations were defined as $\geq 3\times$ upper limit of normal or $\geq 100$ U/L. ALT indicates alanine aminotransferase; AST, aspartate aminotransferase; MACE, major adverse cardiovascular events; HCC, hepatocellular carcinoma; CKD, chronic kidney disease; INR, international normalized ratio; IR, incidence rate; SV, social vulnerability.

SV was also associated with higher risks of HIV viremia (HR, 2.05; 95% CI, 1.71-2.46) and CD4 count <200 cells/µL (HR, 1.58; 95% CI, 1.28-1.95). Extrahepatic organ outcomes were elevated with SV, including MACE (HR, 1.47; 95% CI, 1.22-1.77), ischemic heart disease (HR, 1.61; 95% CI, 1.29-2.02), cerebrovascular disease (HR, 1.67; 95% CI, 1.30-2.15), heart failure (HR, 1.99; 95% CI, 1.51-2.62), CKD (HR, 1.49; 95% CI, 1.20-1.86), and diabetes (HR, 1.25; 95% CI, 1.04-1.49) (**Table 2**).

### Outcomes by Social Vulnerability Intensity

Associations generally strengthened with greater SV intensity for most nonmortality outcomes (**Table 3**). Mortality risk was higher with single-domain than multidomain vulnerability (HR, 1.99; 95% CI, 1.61-2.45 vs. HR, 1.61; 95% CI, 1.19-2.17). However, multidomain SV showed stronger associations than single-domain SV for most hepatic, virologic, liver synthetic function, HIV disease control, and extrahepatic organ outcomes. The liver composite increased from HR 1.41 with single-domain vulnerability to HR 2.49 with multidomain vulnerability, with similar gradients for hepatic decompensation (HR, 1.76 vs. 2.62), HBV DNA ≥2,000 IU/mL (HR, 1.51 vs. 1.99), significant ALT elevation (HR, 1.35 vs. 1.76), and significant AST elevation (HR, 1.37 vs. 1.85). Multidomain gradients were also observed for thrombocytopenia, hypoalbuminemia, coagulopathy, HIV viremia, CD4 count <200 cells/µL, MACE, heart failure, cerebrovascular disease, CKD, and diabetes. Full single-domain and multidomain analyses are shown in **Supplementary Tables S3 and S4**.

**Table 3.** Outcomes by Social Vulnerability Intensity in Propensity Score-Matched Adults With HIV/HBV Coinfection.

| Outcome | 1-domain SV<br>(N=2,944) |  | ≥2-domain SV<br>(N=1,490) |  |
| --- | --- | --- | --- | --- |
|  | HR (95% CI) | p Value | HR (95% CI) | p Value |
| **Survival** |  |  |  |  |
| All-cause mortality | 1.99 (1.61–2.45) | **<0.001** | 1.61 (1.19–2.17) | **0.002** |
| **Liver disease and HBV viremia** |  |  |  |  |
| Liver composite | 1.41 (1.02-1.93) | **0.035** | 2.49 (1.62-3.85) | **<0.001** |
| Hepatic decompensation | 1.76 (1.17-2.65) | **0.006** | 2.62 (1.53-4.48) | **<0.001** |
| **HBV viremia and liver inflammation** |  |  |  |  |
| HBV DNA ≥2,000 IU/mL | 1.51 (1.10-2.08) | **0.010** | 1.99 (1.36-2.91) | **<0.001** |
| Significant ALT elevation (≥100 IU/L) | 1.35 (1.03–1.77) | **0.030** | 1.76 (1.30-2.40) | **<0.001** |
| Significant AST elevation (≥100 IU/L) | 1.37 (1.10–1.71) | **0.006** | 1.85 (1.40–2.44) | **<0.001** |
| **Liver synthetic function** |  |  |  |  |
| Thrombocytopenia (platelet count <150 ×10<sup>9</sup>/L) | 1.25 (0.99-1.57) | 0.066 | 1.85 (1.39-2.48) | **<0.001** |
| Hypoalbuminemia (albumin <3.5 g/dL) | 1.45 (1.17-1.81) | **0.001** | 2.32 (1.76-3.05) | **<0.001** |
| Coagulopathy (INR >1.5) | 1.48 (1.17–1.88) | **0.001** | 2.48 (1.84–3.35) | **<0.001** |
| **HIV disease control** |  |  |  |  |
| HIV RNA >50 copies/mL | 1.87 (1.50-2.33) | **<0.001** | 2.05 (1.54-2.74) | **<0.001** |
| CD4 count <200 cells/μL | 1.42 (1.09-1.85) | **0.010** | 2.09 (1.51-2.88) | **<0.001** |
| **Extrahepatic outcomes** |  |  |  |  |
| MACE | 1.47 (1.17-1.84) | **0.001** | 1.68 (1.25-2.27) | **0.001** |
| Ischemic heart disease | 1.52 (1.17-1.98) | **0.002** | 1.54 (1.10-2.16) | **0.011** |
| Heart failure | 1.56 (1.12-2.19) | **0.009** | 2.17 (1.45-3.25) | **<0.001** |
| Cerebrovascular disease | 1.77 (1.27-2.47) | **0.001** | 2.40 (1.65-3.49) | **<0.001** |
| CKD | 1.27 (0.99-1.65) | 0.064 | 1.72 (1.22-2.42) | **0.002** |
| Diabetes | 1.35 (1.08-1.68) | **0.008** | 1.68 (1.26-2.23) | **<0.001** |

Hazard ratios compare adults with HIV/HBV coinfection with 1-domain or ≥2-domain social vulnerability with propensity score-matched adults with HIV/HBV coinfection and no documented social vulnerability. N indicates the total matched cohort size for each comparison; outcome-specific denominators may vary after exclusion of individuals with pre-follow-up evidence of the corresponding outcome. Significant ALT and AST elevations were defined as ≥3× upper limit of normal or ≥100 U/L. ALT indicates alanine aminotransferase; AST, aspartate aminotransferase; MACE, major adverse cardiovascular events; CKD, chronic kidney disease; INR, international normalized ratio.

### Outcomes by Isolated Social Vulnerability Domain

Isolated-domain analyses suggested that individual SV domains marked distinct patterns of risk (**Table 4**). Healthcare access and engagement vulnerability showed the broadest pattern of association. Compared with adults with no documented SV, those with isolated healthcare vulnerability had more than 3-fold higher mortality (HR, 3.23; 95% CI, 2.51-4.17) and higher risks of the liver composite, HBV viremia, HIV viremia, CD4 count <200 cells/µL, MACE, CKD, and diabetes. Material vulnerability showed a more selective pattern. This domain was associated with higher mortality (HR, 2.11; 95% CI, 1.17-3.81), HIV viremia (HR, 2.09; 95% CI, 1.17-3.73), and MACE, but not clearly with the liver composite, HBV viremia, CD4 count <200 cells/µL, CKD, or diabetes. Interpersonal/psychosocial vulnerability was associated with MACE (HR, 1.76; 95% CI, 1.06-2.94), but not with mortality, the liver composite, HBV viremia, HIV viremia, CKD, or diabetes.

**Table 4.** Outcomes by Isolated Social Vulnerability Domain in Propensity Score-Matched Adults With HIV/HBV Coinfection.

| Outcomes | Material vulnerability<br>(N=568) |  | Healthcare vulnerability<br>(N=1,780) |  | Interpersonal/Psychosocial<br>vulnerability (N=614) |  |
| --- | --- | --- | --- | --- | --- | --- |
|  | HR (95% CI) | p Value | HR (95% CI) | p Value | HR (95% CI) | p Value |
| **Survival** |  |  |  |  |  |  |
| All-cause mortality | 2.11 (1.17-3.81) | **0.012** | 3.23 (2.51-4.17) | **<0.001** | 1.31 (0.77-2.21) | 0.317 |
| **Liver disease and HBV**<br>**viremia** |  |  |  |  |  |  |
| Liver composite | 0.68 (0.41-1.14) | 0.138 | 1.57 (1.07-2.29) | **0.020** | 0.95 (0.61-1.48) | 0.826 |
| HBV DNA $\geq 2,000$ IU/mL | 1.55 (0.70-3.42) | 0.274 | 1.49 (1.02-2.18) | **0.037** | 1.04 (0.46-2.36) | 0.934 |
| **HIV disease control** |  |  |  |  |  |  |
| HIV RNA >50 copies/mL | 2.09 (1.17-3.73) | **0.011** | 2.15 (1.62-2.85) | **<0.001** | 1.05 (0.64-1.74) | 0.840 |
| CD4 count <200 cells/μL | 0.66 (0.35-1.26) | 0.202 | 1.57 (1.14-2.16) | **0.005** | 0.50 (0.26-1.01) | 0.068 |
| **Extrahepatic outcomes** |  |  |  |  |  |  |
| MACE | 1.7 (1.07-2.88) | **0.024** | 1.43 (1.05-1.95) | **0.023** | 1.76 (1.06-2.94) | **0.027** |
| CKD | 0.98 (0.53-1.82) | 0.942 | 1.71 (1.20-2.44) | **0.003** | 1.21 (0.77-1.89) | 0.412 |
| Diabetes | 1.04 (0.58-1.86) | 0.903 | 1.34 (1.03-1.75) | **0.031** | 1.01 (0.65-1.58) | 0.953 |

Hazard ratios compare adults with HIV/HBV coinfection with isolated single-domain social vulnerability with propensity score-matched adults with HIV/HBV coinfection and no documented social vulnerability. N indicates the total matched cohort size for each comparison; outcome-specific denominators may vary after exclusion of individuals with pre-follow-up evidence of the corresponding outcome. Interpersonal and psychosocial vulnerability domains were combined because of limited sample size. MACE indicates major adverse cardiovascular events; CKD, chronic kidney disease.

### Sensitivity Analyses

Findings were robust across prespecified sensitivity analyses. In the key healthcare utilization-adjusted sensitivity analysis, associations persisted after accounting for ambulatory, emergency department, and hospitalization-related healthcare use (**Supplementary Table S5**). SV remained associated with mortality (HR, 1.91; 95% CI, 1.56-2.34), liver composite events (HR, 1.44; 95% CI, 1.19-1.74), hepatic decompensation (HR, 1.97; 95% CI, 1.38-2.82), hepatic failure (HR, 2.66; 95% CI, 1.62-4.36), HBV DNA ≥2,000 IU/mL (HR, 1.70; 95% CI, 1.28-2.26), and HIV viremia (HR, 1.73; 95% CI, 1.44-2.07). Cardiovascular outcomes, CKD, and diabetes also remained elevated. These findings indicate that the observed associations were not explained solely by differential baseline healthcare contact.

In additional sensitivity analyses, findings remained consistent in the 6-month lag analysis, including persistent associations with mortality, liver composite events, hepatic decompensation or failure, HBV DNA ≥2,000 IU/mL, HIV viremia, CD4 count <200 cells/µL, and cardiovascular outcomes (**Supplementary Table S6**). Results were also similar after expanded matching for CD4 count, HIV RNA, HBV DNA, and ART, including persistent associations with mortality, liver composite events, hepatic decompensation or failure, HBV viremia, HIV viremia, MACE, CKD, and diabetes (**Supplementary Table S7**).

## DISCUSSION

In this large propensity score-matched cohort of adults with HIV/HBV coinfection in the United States, documented SV was associated with higher mortality and worse hepatic, virologic, and extrahepatic organ outcomes. These associations persisted across multiple sensitivity analyses, including the key healthcare utilization-adjusted analysis accounting for ambulatory visits, emergency department visits, and hospitalizations. This sensitivity analysis directly addresses the concern that adults with more healthcare contact may have more opportunities for both SV documentation and outcome ascertainment. Risk also varied by SV intensity: multidomain vulnerability was associated with larger hazard ratios than single-domain vulnerability for most nonmortality outcomes, supporting a graded relationship. Isolated-domain analyses further suggested that different domains may correspond to distinct clinical risk profiles, particularly for hepatic, virologic, and extrahepatic organ outcomes. These findings extend the social determinants of health literature by demonstrating that dynamic, multidimensional SV is associated with clinically consequential outcomes in HIV/HBV coinfection [29].

The 2-fold higher mortality associated with SV is consistent with, though larger than, prior estimates from HIV cohorts. In the ANRS CO8 APROCO-COPILOTE cohort, SV was associated with a 20% higher mortality risk among people with HIV receiving ART [33], and other studies have similarly linked housing instability and food insecurity to increased mortality risk among people with HIV [34,35]. The larger effect size observed here may reflect the compounded clinical demands of dual infection: HIV/HBV coinfection requires uninterrupted HBV-active ART adherence, HCC surveillance, and liver disease monitoring, creating multiple points at which social disruption can translate into clinical harm [22,34–36]. The association of SV with hepatic decompensation and hepatic failure, but not clearly with cirrhosis or HCC, suggests that social disruption may more readily precipitate clinical destabilization among adults with underlying liver vulnerability than produce new cirrhosis or HCC during follow-up. The null findings for cirrhosis and HCC may also reflect the longer natural history of these outcomes, which evolve over years to decades and may be less readily captured within the observation window [4,30,31,37]. Mechanistically, ART or tenofovir interruptions can precipitate HBV DNA rebound, hepatitis flares, and HIV-related morbidity, particularly when HBV monitoring is infrequent [30,31,36]. Thus, the higher risks of HIV and HBV viremia, transaminase elevations, and impaired liver function observed in the SV group support treatment continuity as a plausible pathway linking SV to liver-related morbidity.

Beyond liver disease, HIV/HBV coinfection may also contribute to systemic morbidity through persistent antigenic stimulation, immune activation, microbial translocation, and gut-liver immune signaling [38–40]. Simultaneously, chronic stress from SV may become biologically embedded through hypothalamic-pituitary-adrenal axis dysregulation and allostatic load, amplifying systemic inflammation, endothelial dysfunction, insulin resistance, and renal injury [41,42]. Our finding of elevated CRP and ESR among adults with SV supports systemic inflammation as a potential downstream pathway. Thus, the excess cardiometabolic and renal burden observed here may reflect the convergence of viral persistence, care disruption, and stress biology rather than hepatic disease alone [22,36,38–42]. Consistent with this model, SV was also associated with worse HIV disease control, including a 2-fold higher risk of HIV viremia and a 58% higher risk of CD4 count <200 cells/µL. These findings align with Medical Monitoring Project data showing that cumulative social and economic disadvantage is associated with missed visits and lower viral suppression among people with HIV in a dose-dependent fashion [43]. In HIV/HBV coinfection, disrupted viral control may therefore function as both a marker of care instability and a mediator of downstream inflammatory, cardiovascular, metabolic, and renal risk [41,43].

The isolated-domain and intensity analyses provided an additional layer of support for interpreting SV as a multidimensional clinical risk phenotype in HIV/HBV coinfection [26,29]. In isolated-domain analyses, healthcare access and engagement vulnerability was most strongly associated with mortality and was the most consistent correlate of liver disease, hepatic decompensation, HBV viremia, and HIV viremia. This domain-specific pattern aligns with the clinical demands of HIV/HBV care, which requires uninterrupted HBV-active ART and regular monitoring, creating multiple points at which care disruption can threaten viral and hepatic control [22,30,31,36]. Material vulnerability showed a more selective association with mortality, HIV viremia, and MACE, suggesting that housing, food, transportation, and economic instability may contribute to clinical risk through broader pathways of resource deprivation and care instability [13,18,19,34,35]. Interpersonal/psychosocial vulnerability was less clearly associated with liver or virologic outcomes in isolated-domain analyses but remained relevant to cardiovascular risk. These domain-specific patterns indicate that SV should not be treated as a single undifferentiated exposure [26,29].

Our findings may have several clinical and policy implications. First, they support integrating systematic SV assessment into HIV/HBV care. Second, the graded relationship between SV and risk accumulation suggests that identifying the number of active SV domains, not merely the presence or absence of any one SV indicator, may improve clinical risk stratification [23,26–29]. Third, domain-specific patterns suggest that interventions should be tailored: healthcare access and engagement vulnerability should trigger urgent care re-engagement, insurance navigation, pharmacy support, and monitoring recovery; material vulnerability should prompt linkage to housing, food, transportation, and financial assistance; and multidomain vulnerability should activate comprehensive case management. These intervention priorities align with evidence that social determinants of health-focused hepatitis B interventions can improve care-related outcomes [17].

Several limitations should be considered alongside the study’s strengths. SV was identified using diagnosis codes, including Z codes, which are underused in clinical documentation, with reported use in only approximately 1% to 2% of medical records [44]. This likely underestimated SV and may have selected for individuals with more clinically visible social needs. Care disengagement and nonadherence codes identify documented care engagement problems but do not specify whether the underlying reason was structural, behavioral, clinical, or undocumented. TriNetX also does not fully capture people who are uninsured, receive care outside participating networks, or are completely disengaged from care. Differential healthcare contact may influence SV documentation and outcome ascertainment; however, the key healthcare utilization-adjusted sensitivity analysis showed that associations persisted after accounting for ambulatory, emergency department, and hospitalization-related healthcare utilization. Residual confounding remains possible despite propensity score matching, including by unmeasured treatment adherence, ART regimen details, hepatitis D coinfection, alcohol use severity, and social risk severity. The observational design precludes causal inference, and SV may be both a driver and consequence of clinical deterioration. Some subgroup analyses were limited by small event counts. These limitations are balanced by the large, geographically diverse U.S. cohort; prespecified four-domain framework; evaluation of outcomes across hepatic, virologic, cardiovascular, renal, and metabolic domains; robust propensity score matching; and multiple sensitivity analyses.

In summary, in this large U.S. cohort of adults with HIV/HBV coinfection, documented SV was associated with higher mortality, hepatic decompensation, hepatic failure, impaired HIV and HBV viral control, and extrahepatic organ morbidity. These associations persisted in the key healthcare utilization-adjusted sensitivity analysis, suggesting that findings were not explained solely by differential baseline healthcare contact. Risk increased with SV intensity, with multidomain vulnerability identifying a higher-risk phenotype for most nonmortality outcomes. Isolated-domain analyses showed that healthcare access and engagement vulnerability was the strongest domain for mortality, liver, and virologic outcomes. These findings support integrating dynamic, multidimensional SV assessment into HIV/HBV care and suggest that domain-targeted interventions may be needed to reduce excess morbidity and mortality among socially vulnerable adults with HIV/HBV coinfection.

## Data Availability

All data produced in the present work are contained in the manuscript

## AUTHOR CONTRIBUTIONS

GAY conceptualized the study with input from KD. GAY curated the data and performed the statistical analyses. GAY, TC, AD, AA, AMM, CC, MLF, HNK, IO, and KD contributed to the development of the methods, analytic approach, and interpretation of findings. GAY secured funding and oversaw project administration. GAY drafted the original manuscript. All authors critically reviewed and revised the manuscript for important intellectual content and approved the final version. All authors had full access to the data. GAY and KD had final responsibility for the decision to submit the manuscript for publication.

## DATA SHARING

Deidentified individual participant data that underlie the results were extracted from TriNetX, a federated national health research network with data sourced from 110 health care organizations within the United States with waiver from WCG IRB.

